# Incidence of Acute Watery Diarrhea among Internally Displaced People in Burhakaba Camps, Bay Region, Somalia

**DOI:** 10.1101/2023.09.10.23295320

**Authors:** Mohamed Jayte, Maryan Dahir

**Affiliations:** Internal Medicine Department of Kampala International University Teaching Hospital., Kampala, Uganda; Public Health Department at Somali International University

**Keywords:** Acute watery diarrhea, incidence, risk factors, internally displaced persons, Somalia

## Abstract

**Background:** Acute watery diarrhea (AWD) is a major public health issue among internally displaced persons (IDPs) in Somalia due to inadequate water, sanitation and hygiene (WASH) infrastructure and services, in crowded camps. This study aimed to determine the incidence and associated factors of AWD in IDP camps in Burhakaba, Bay region.

**Methods:** A cross-sectional survey was conducted among 318 IDPs across 6 camps in Burhakaba town. Structured interviews collected data on demographics 7-day episodes of AWD, water sources, latrine access, hygiene practices, and food consumption. AWD incidence was calculated and bivariate analysis identified associated factors.

**Results:** The overall AWD incidence was 16.7%. Children under 5 had the highest incidence, at 9.8% among under 1 year and 8.9% among 1-5 years. Use of open wells (p=0.026), lack of latrines (p=0.017), poor hand-washing practices (p=0.002), and reduced food intake (p<0.001) were significantly associated with higher AWD risk.

**Conclusions:** Targeted intervention to improve water quality, sanitation coverage, hygiene practices, and food security are urgently needed to reduce the high AWD burden among IDPs in Burhakaba.

## Introduction

Acute watery diarrhea (AWD) remains a leading cause of morbidity and mortality, especially among children under 5 years, in low income countries (1). AWD is typically caused by ingestion of contaminated food or water infected with viral, bacterial or protozoal pathogens like rotavirus, Vibrio cholerae, Shigella spp. and Cryptosporidium spp. among others (2). Disease transmission is facilitated by poor water quality, sanitation and hygiene (WASH) conditions, overcrowding, and malnutrition (3). Internally displaced persons (IDPs) living in crowded camp conditions with inadequate WASH facilities and food insecurity are especially vulnerable to AWD outbreaks (4).

In Somalia, recurrent AWD outbreaks have occurred among IDPs since the 2011 famine (5). The 2017 AWD outbreak in Somalia resulted in 78,783 suspected cases and 1,159 deaths (Case Fatality Rate 1.5%) between January 1 and July 23, 2017 (6). Bay region was the second most affected area in Somalia, with 13,370 cases and 151 deaths (7). Persistent risk factors in IDP settlements provide conditions ripe for repeated AWD outbreaks.

This study aimed to determine the incidence and associated factors of AWD among IDPs in Burhakaba camps, Bay region, Somalia, in order to guide future AWD prevention and control efforts. Specific objectives were to estimate the overall and age-specific incidence of AWD and Identify risk factors for AWD associated with water sources, sanitation, hygiene practices and food security.

## Methods

### Study design and setting

We conducted a cross-sectional community-based survey in May 2020 among IDPs in 6 camps (Buurhakaba, Isneeb, Magaalijecel, Buulinasiib, Awyaaye and Daarusalaam) in Burhakaba town, Bay region, southwestern Somalia. Burhakaba town hosts approximately five thousand eight hundred IDP households (35,000 individuals) across several crowded camps with makeshift shelters (8). The camps have limited clean water, latrines, health services and food supplies (8).

### Study population and sample size

The study population was IDPs of all ages residing in the Burhakaba camps. Sample size was calculated using a 50% expected AWD prevalence (no recent data), 5% precision and 95% confidence level, giving a minimum sample of 384. An additional 10% was added to account for non-response, giving a final sample size of 422. This was proportionately allocated to the 6 camps based on available population data (8). Households were randomly selected and all members were surveyed.

### Data collection

A structured questionnaire adapted from WHO (9) was used to collect data on demographics, AWD episodes, water sources, sanitation, hygiene practices and food consumption. Interviews were conducted with the head of household or spouse. For children under 12 years, responses were given by the primary caregiver.

### Data analysis

Data were entered into EpiData v3.1 and analyzed using EpiData Analysis v2.2.2.183. Frequencies and proportions were calculated. AWD incidence was calculated by dividing the number of AWD cases by the total population surveyed. Prevalence was also determined by age group. Bivariate analysis using chi-square test was done to assess associations between AWD and explanatory variables at p<0.05 significance.

## Results

### Sociodemographic characteristics

A total of 318 IDPs were surveyed, with 54% females (Table 1). The mean household size was 5.4 persons. Children under 5 years comprised 37% of the sample size. The majority (62%) had no formal education.

**Table 1.**
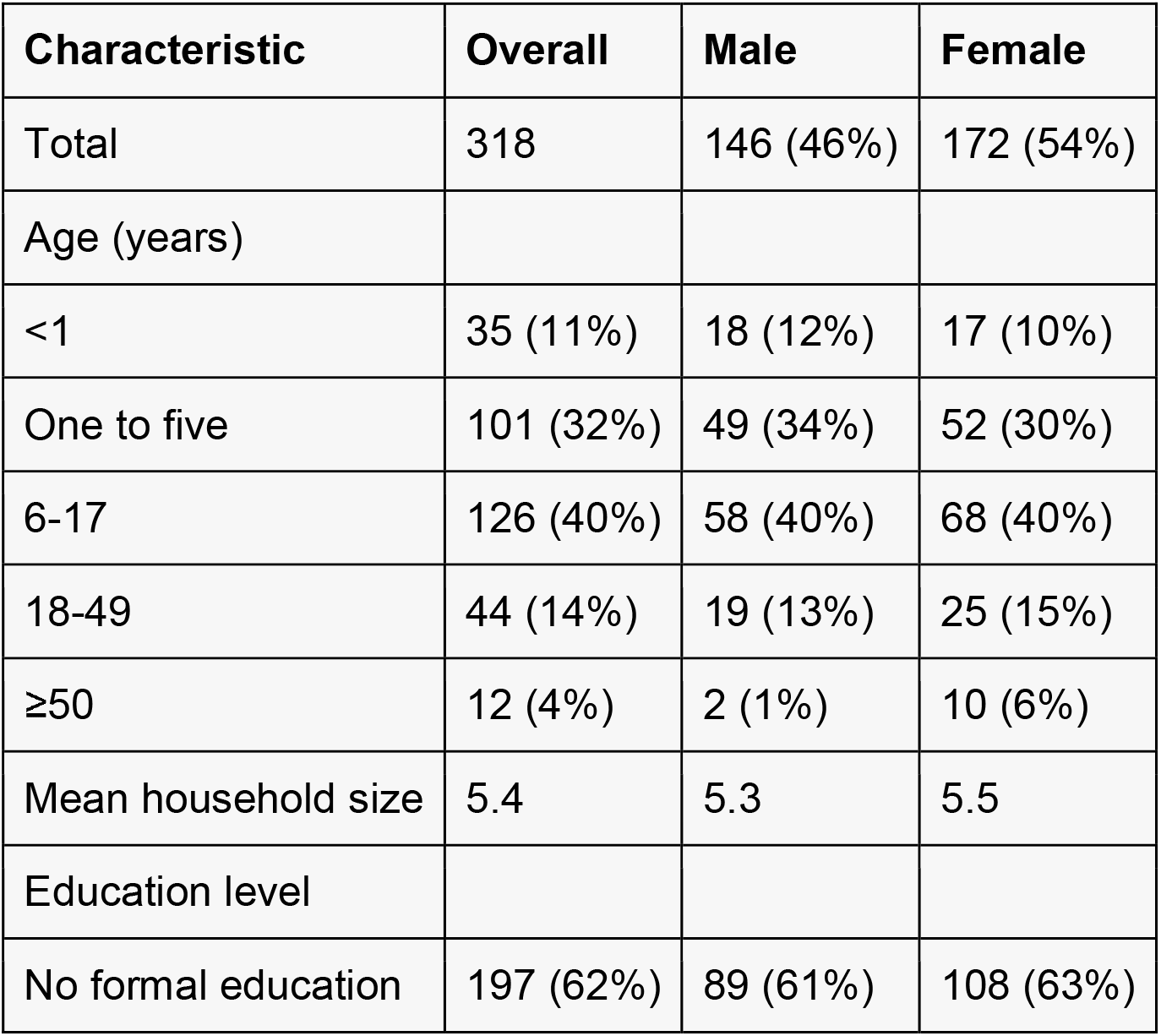
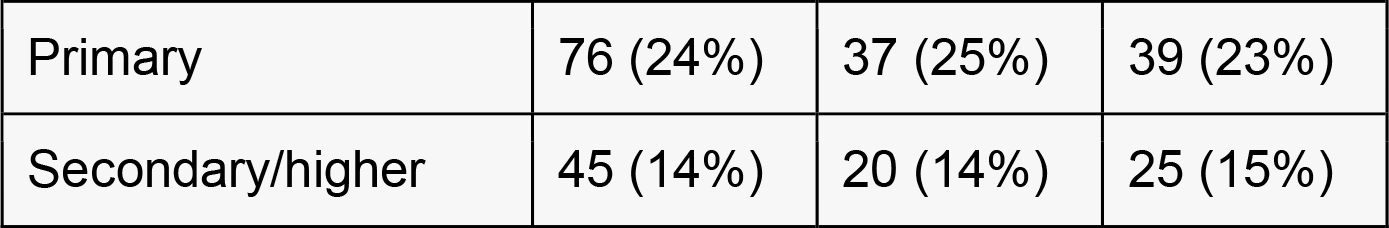
Demographics (n=318)

### AWD Incidence

The overall AWD incidence among the surveyed population was 16.7% (Table 2). Incidence was highest in Daarusalaam camp, (48%) followed by Buulinasiib (28%). Children under 5 years had the highest age-specific incidence of 9.8% among under 1 year olds and 8.9% in 1-5 year olds. The incidence decreased with increasing participant age, with the lowest of 2.2% in those over 50 years.

Table presenting the AWD Incidence by camp and by age group

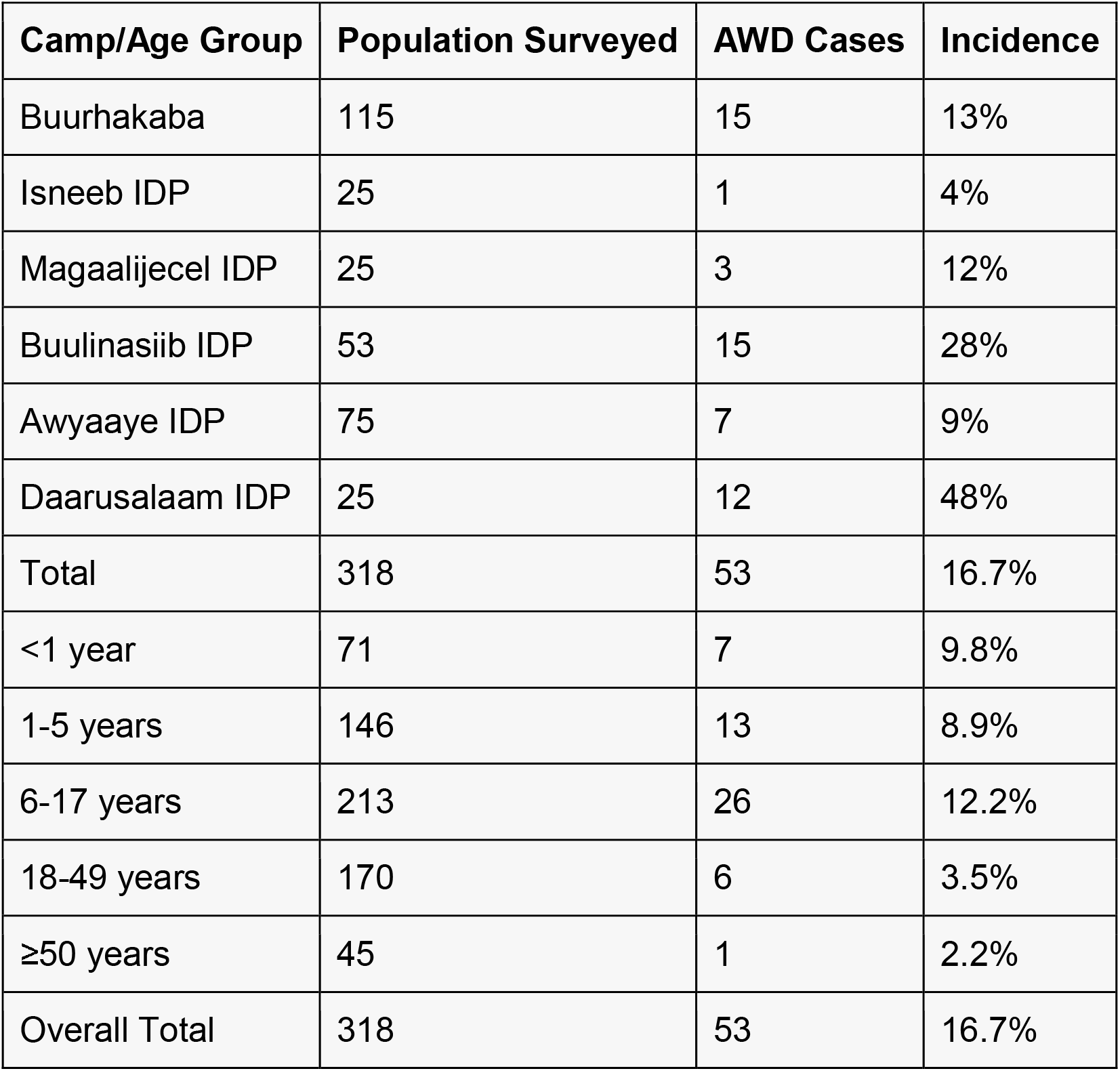

| Camp/Age Group | Population Surveyed | AWD Cases | Incidence |
| --- | --- | --- | --- |
| Buurhakaba | 115 | 15 | 13% |
| Isneeb IDP | 25 | 1 | 4% |
| Magaalijecel IDP | 25 | 3 | 12% |
| Buulinasiib IDP | 53 | 15 | 28% |
| Awyaaye IDP | 75 | 7 | 9% |
| Daarusalaam IDP | 25 | 12 | 48% |
| Total | 318 | 53 | 16.7% |
| <1 year | 71 | 7 | 9.8% |
| 1-5 years | 146 | 13 | 8.9% |
| 6-17 years | 213 | 26 | 12.2% |
| 18-49 years | 170 | 6 | 3.5% |
| ≥50 years | 45 | 1 | 2.2% |
| Overall Total | 318 | 53 | 16.7% |

### Risk factors for AWD

Most respondents (61%) reported using open wells and boreholes as water sources, which are prone to contamination (Table 3). Only 30% had access to latrines, with most (62%) practicing open defecation. Less than half (41%) reported always washing hands before eating and after defecation. The majority (71%) reported reduced food intake in the past week due to shortages. Bivariate analysis showed significant associations between AWD and use of open wells (p=0.026), lack of latrine access (p=0.017), inconsistent hand-washing (p=0.002), and reduced food intake (p<0.001).

**Table 3.**
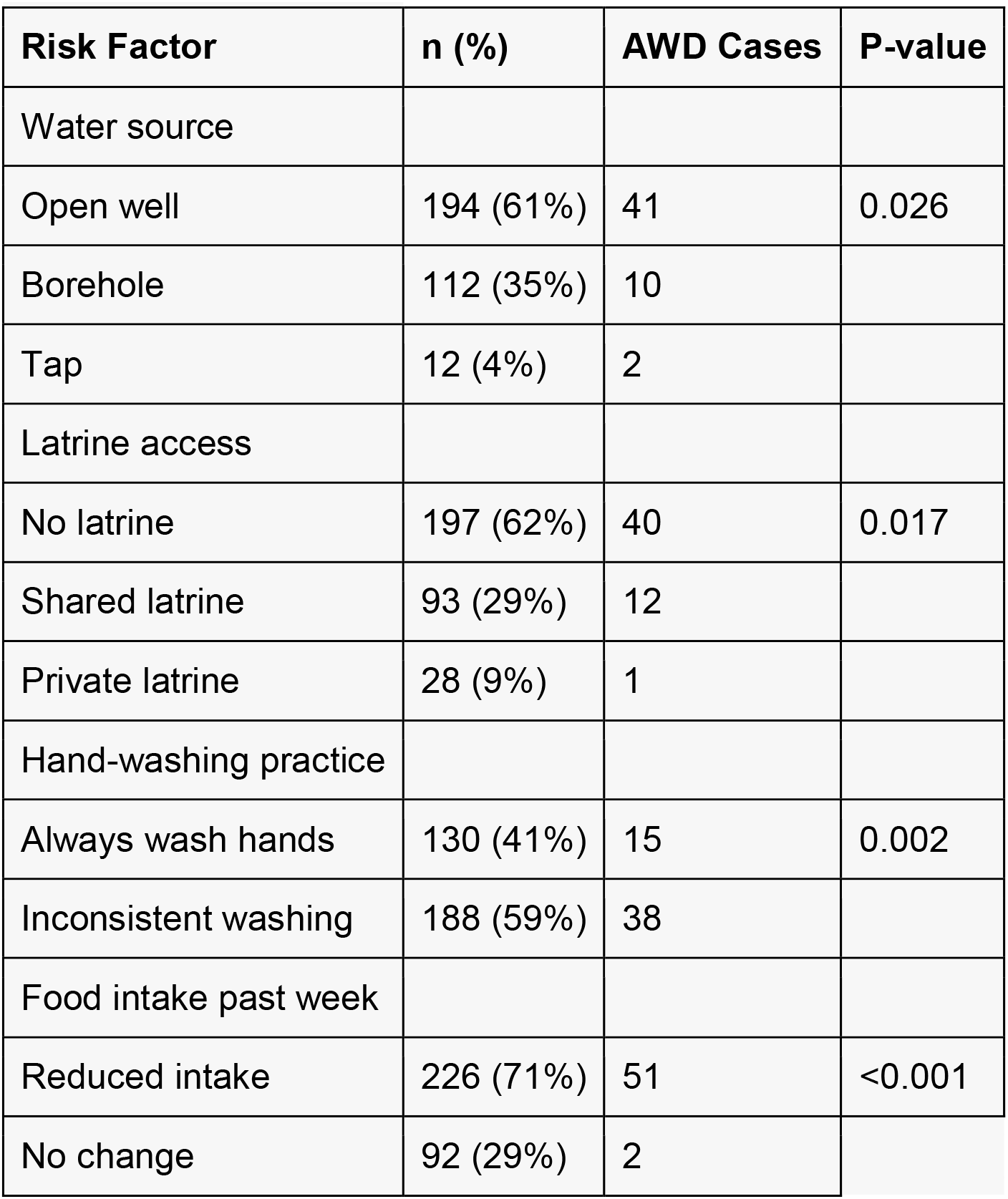
Risk factors for AWD.

## Discussion

This study found a high AWD incidence of 16.7% among IDPs in Burhakaba camps, Bay region, Somalia. The IDP camps have persistently high AWD transmission due to crowding, inadequate safe water and sanitation facilities, poor hygiene practices, and widespread food insecurity. Children under five years bore the greatest burden with incidence over 9%, likely due to their greater susceptibility to dehydration and malnutrition.

The significant association between AWD and use of open wells indicates water contamination was a likely contributor to disease spread. Most open wells in Somalia are unprotected and prone to runoff contamination with human and animal waste (10). Proper well protection such as lining, covers, drainage channels and fencing should be implemented in IDP camps to prevent water contamination (11). Household water treatment like chlorination before use is also recommended.

Lack of latrines was significantly associated with higher AWD risk as most IDPs practice open defecation. This facilitates contamination of water and food by fecal-oral transmission of pathogens (12). The Construction of adequate numbers of improved pit latrines strategically located for safety is crucial to control open defecation and safe disposal of child and adult feces (13). Proper use and maintenance through hygiene promotion must accompany latrine provision.

Inconsistent hand-washing before eating and after defecation or cleaning child feces was an AWD risk factor. Hand-washing with soap removes pathogens responsible for diarrheal diseases (14). to “Constructing hand-washing stations at latrines, food preparation areas, and household entrances combined with hygiene education are needed. Soap provision may encourage hand-washing practice.

Reduced food intake due to shortages was significantly associated with AWD. Malnutrition impairs immune function and mucosal integrity, increasing susceptibility to enteric infections and progression to life-threatening diarrhea (15). Improved food security through supplementary feeding programs and livelihood support interventions can mitigate this risk factor for vulnerable groups like children under 5 years, pregnant women, elderly and disabled people (16).

This study had some limitations. First, self-reported AWD episodes may be affected by recall bias. Second, loose stool samples were not collected for laboratory analysis to confirm etiologic agents. However, most AWD outbreaks in Somalia have been attributed to Vibrio cholerae O1 Inaba confirming the clinical case definitions are valid for cholera surveillance (6). Despite these limitations, the study found a high AWD burden in Burhakaba IDP camps associated with poor WASH conditions and food insecurity, reflecting common challenges across Somalia (5,10).

## Conclusion

This study found a high AWD incidence among IDPs in Burhakaba camps, Bay region, Somalia, with the highest burden in children under 5 years. Contaminated water sources, inadequate sanitation, poor hygiene practices and food insecurity were significantly associated with increased risk. Priority interventions include improving water quality, increasing latrine coverage, promoting hand-washing with soap, and supplementary feeding programs targeting malnourished groups. Coordinated efforts between humanitarian agencies and local authorities are needed to upgrade WASH infrastructure, provide hygiene education, and strengthen outbreak preparedness in these vulnerable IDP camps. Addressing these systemic issues and enhancing regular surveillance can help reduce the unacceptable AWD disease burden among displaced populations in Somalia.

## Data Availability

All data produced in the present study are available upon reasonable request to the authors

## Ethics Considerations

Ethical approval was obtained from the Ministry of Health Somalia. Informed verbal consent was obtained from all participants prior to the interviews. Sick individuals were referred to the nearest health facility for treatment.

## Access to Research Data

The datasets used and analyzed for this research are accessible by contacting the study’s corresponding author and submitting a reasonable request.

## Abbreviations

AWD: Acute watery diarrhea
IDP: Internally displaced person
WASH: Water, sanitation and hygiene
UNICEF: United Nations Children’s Fund
WHO: World Health Organization
CFR: Case fatality rate
EpiData: Epidemiological data software
SPSS: Statistical Package for the Social Sciences

